# An analysis of humanitarian and health aid harmonisation over a decade (2011-2019) of the Syrian conflict

**DOI:** 10.1101/2024.04.17.24305968

**Authors:** Munzer Alkhalil, Abdulkarim Ekzayez, Kristen Meagher, Maher Alaref, Rim Turkmani, Aula Abbara, Zedoun Alzoubi, Nassim El Achi, Preeti Patel

## Abstract

**Introduction:** Aid harmonisation is a key component of donor efforts to make aid more effective by improving coordination and simplifying and sharing information to avoid duplication. This study evaluates the harmonisation of health and humanitarian aid in Syria during acute humanitarian and health crises from 2011 to 2019.

**Methods:** Data on humanitarian and health aid for Syria between 2011 and 2019 was collected from the OECD’s Creditor Reporting System (CRS) to assess the harmonisation of aid. The data was linked to four key indicators of the conflict: the number of IDPs; the number of people in need of humanitarian assistance; the number or frequency of internal movements (displacements) by individuals; and the decline in Syria’s population between 2011 and 2019. This was compared to data from highly fragile states and stable low- and middle-income countries. Four focus group discussions and four key informants’ interviews with humanitarian practitioners were also conducted.

**Results:** The findings suggest that overall aid harmonisation did not occur and importantly did not correlate with increased humanitarian needs. During the first five years, humanitarian and health pooled funds (which endorse aid harmonisation) in Syria were nearly entirely absent, far less than those in low- and middle-income countries and highly fragile states. However, from 2016 to 2019, a visible surge in humanitarian pooled funds indicated an increase in the harmonisation of donors’ efforts largely influenced by adopting the Whole-of-Syria approach in 2015 as a positive result of the cross-border UN Security Council resolution in 2014.

**Conclusion:** Harmonisation of aid within the Syrian crisis was found to have little correlation with the crisis parameters and population needs, instead aligning more with donor policies. Assessing fragmentation solely at the donor level is also insufficient. Aid effectiveness should be assessed with the inclusion of community engagement and aid beneficiary perspectives. Harmonisation mechanisms must be disentangled from international politics to improve aid effectiveness. In Syria, this study calls for finding and supporting alternative humanitarian coordination and funding mechanisms that are not dependent on the persistent limitations of the UN Security Council.

**Key Messages:** *What is already known on this topic:* Leading aid agencies must coordinate their policies to manage complex humanitarian and health needs. Yet, the impact of crisis-relevant indicators such as internal displacement trends, and population needs assessments remain underexplored, particularly in the Eastern Mediterranean Region (EMR).

*What this study adds:* Harmonisation of aid in the Syrian crisis did not align with crisis indicators or population needs, but rather with donor policies, creating challenges in the transition from conflict to early recovery. In addition, the applicability of the Paris Declaration on Aid Effectiveness is less relevant in conflict-affected areas, given that it supposes the existence of a legitimate government and leading aid agencies should collaborate to achieve development goals. However, in complex conflicts, governments may have limited control over resources and legitimacy is subsequently questionable.

*How this study might affect research, practice or policy:* This study highlights the need for continuous UN cross-border coordination mechanisms, finding another model than ‘the consent model’ for cross-border humanitarian aid for northwest Syria to protect local communities from ‘aid weaponisation’, and support the newly established Aid Fund for Northern Syria (AFNS), expand its coverage to include northeast Syria and improve its policy to include more development health aid and localisation agenda.

## 1. Introduction

The Syrian crisis is the worst humanitarian crisis of the 21^st^ century [1]. Since protests began in Syria in 2011, an estimated 874,000 people have been killed, directly or indirectly [2]. Of the 22 million people who lived in Syria before 2011, an estimated 13.7 million have become refugees or internally displaced persons (IDPs) [3], [4]. Ninety-eight percent of Syrians now live in extreme poverty, which equates to having less than $1.90 per day per person [5]. The military situation in Syria has seen drastic changes from 2011 to 2019, with the Syrian government struggling to hold power against three de facto local governments that have arisen since 2013 [6]–[13]. The power struggles between these groups have had major consequences for humanitarian and health aid [14], [15].

A UN Security Council Resolution (UNSCR) in 2014 permitted aid delivery via four border crossing points to areas not held by the Syrian government [16]. These four points included Bab al-Salam, Bab al-Hawa, Al Yarubiyah and Al-Ramtha [17]. This led to establishing three humanitarian hubs in Damascus, Syria, Gaziantep, Turkey and Amman, Jordan. In 2019 Russia and China vetoed the extension of the Resolution. An exception was granted to Bab al-Hawa crossing, which continues to allow aid to northwest Syria [18].

Until 2015, the three humanitarian hubs operated independently from each other. But in 2015 donors adopted a Whole of Syria (WoS) approach, bringing together all humanitarian actors working in Syria and neighbouring countries [19]. The aim of this new mechanism, coordinated by the UN, was to increase the effectiveness of the response and reduce risks. This occurred as the complexity of the Syrian context increased as humanitarian actors had to navigate sanctions [20], various non-state actors proscribed as terrorist groups [21], and political red lines drawn by donors [22].

The protracted nature of the conflict [23] has made Syria heavily reliant on emergency humanitarian and, to a lesser extent, development aid, to address local needs [1]. More recently, with the relative decline in hostilities, the international community has considered transitioning the conflict into the early recovery stage [24] while trying to incorporate international initiatives such as the UN’s New Way of Working, comprising the triple nexus and localisation for Syria’s aid support portfolio. Following the World Humanitarian Summit in 2016, the ‘triple nexus’ or the humanitarian-development-peace (HDP) concept has emphasised collaborative efforts for the development, peace and security, and human rights pillars to prioritise prevention, address root causes and support institutions for sustainable peace and development to coherently address vulnerabilities before, during and after crises [25]. This has also emphasised the localisation of aid, reinforcing local ownership and the role of local actors in long term sustainability of capacity strengthening across sectors [26].

As a result, Syria’s main donors and stakeholders have initiated a process of systematising their programmatic interventions to reflect the early recovery phase in the response. Health aid tracking [27] is very useful for most donors and recipients to improve transparency, accountability, timely planning, standardisation, and priority-setting. It is also helpful for learning whether individual donors and recipients have fulfilled their aid commitments, and whether aid was sufficient, targeted those with the greatest needs, effective, and aligned with local strategies.

Aid effectiveness is a key indicator of aid impact and provides local-level analysis in addition to aid tracking. The 2005 Paris Declaration on Aid Effectiveness adopted a practical framework for specific indicators to evaluate aid effectiveness based on five fundamental principles: ownership, alignment, harmonisation, results, and mutual accountability (see Table 1 for definitions) [28].

**Table 1.**
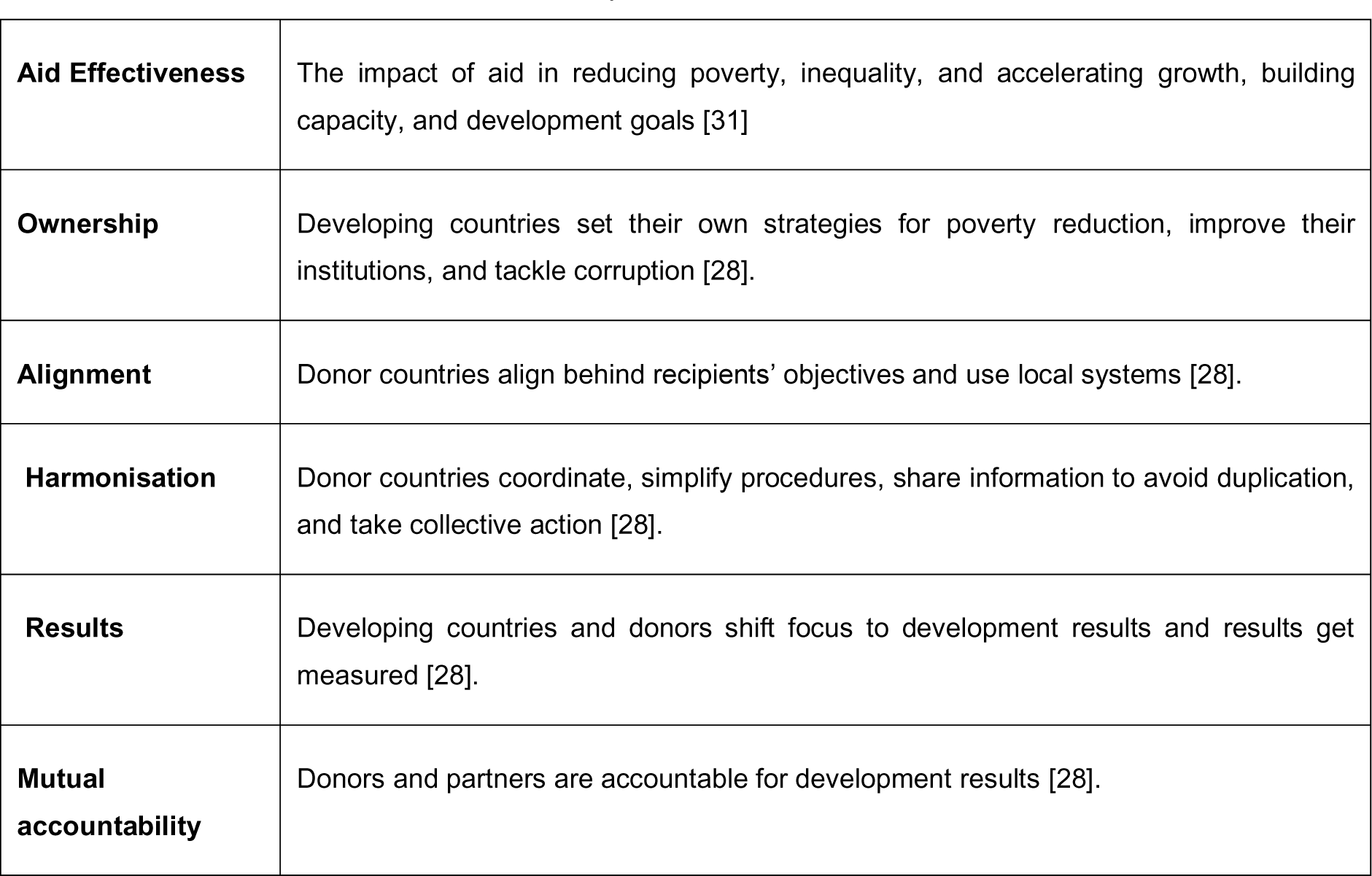
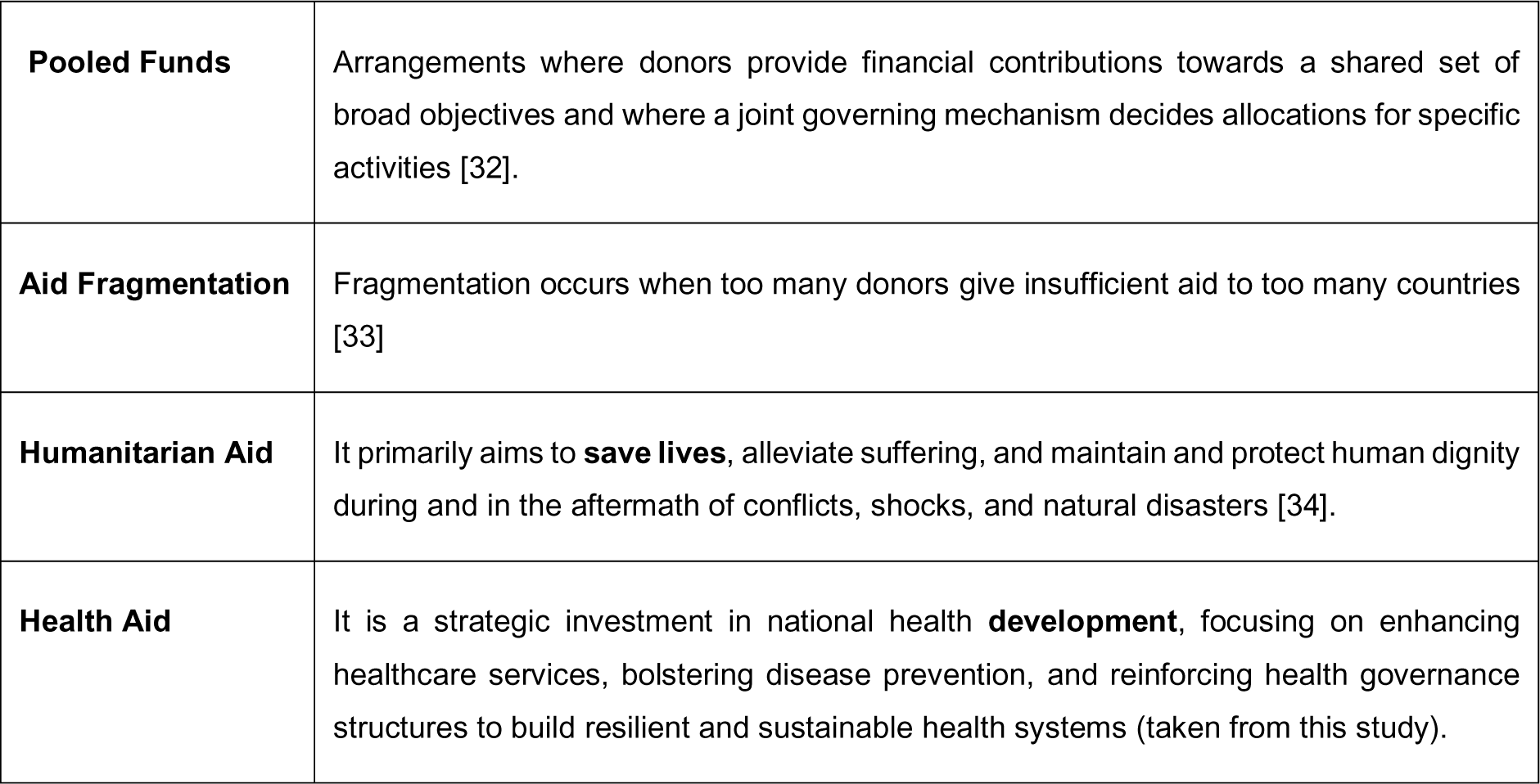
Key Definitions and Terms.

**Table 2.**
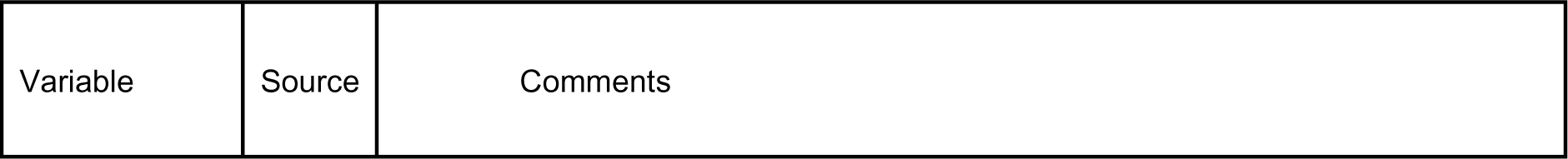
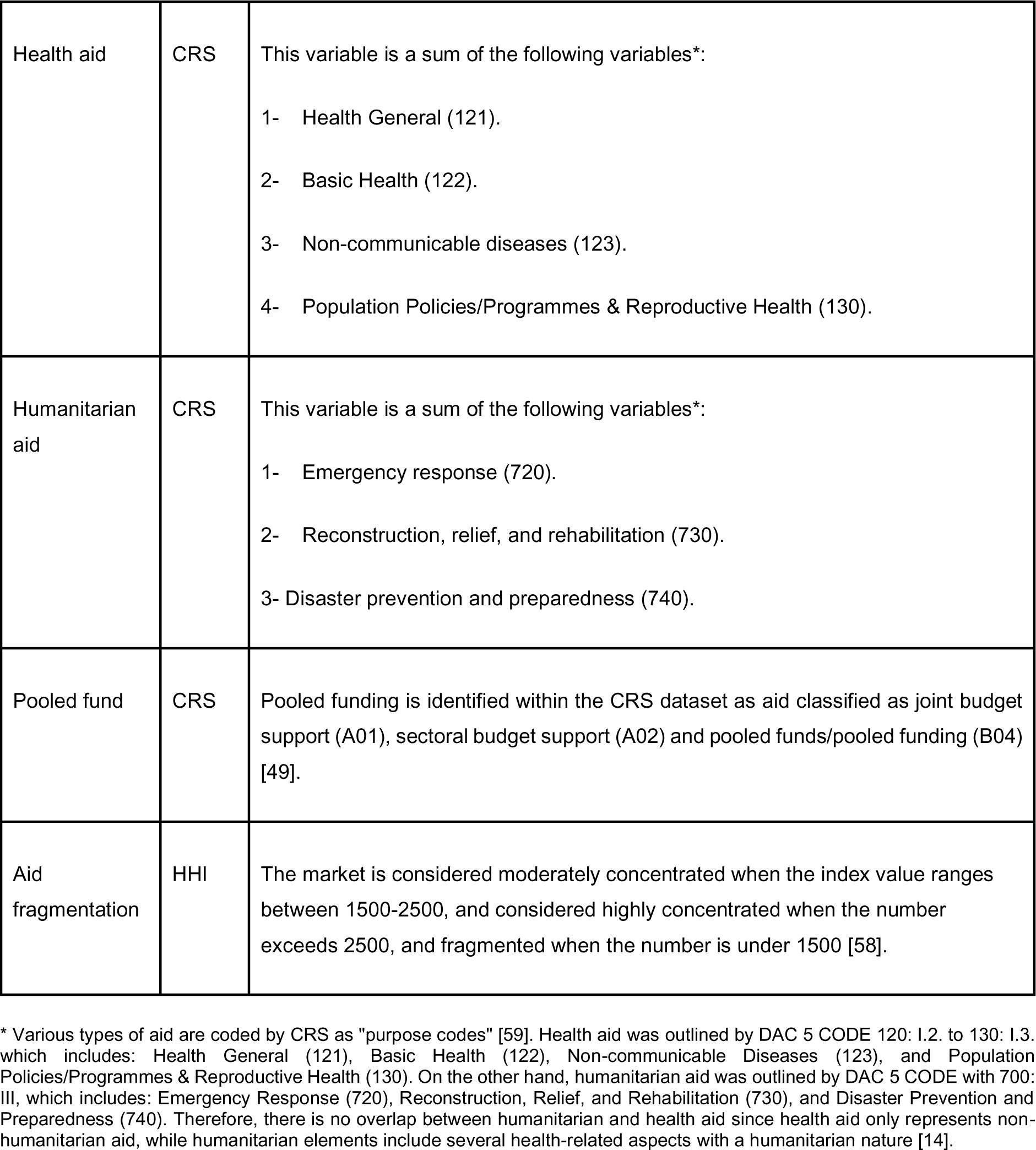
List of variables included in the quantitative analysis.

Aid harmonisation and alignment are crucial in enhancing the efficiency and impact of international aid efforts. Harmonisation ensures that donors collaborate, minimising duplication and thus reducing transaction costs and administrative challenges [29]. Meanwhile, alignment anchors these synchronised endeavours to the recipient country’s intrinsic goals and developmental strategies [30]. This dual approach ensures a synergy where mutual accountability thrives. It empowers and strengthens national systems, ensuring that the benefits of aid are both sustainable and genuinely reflective of the aspirations and requirements of the beneficiary nations, making each aid intervention more potent and meaningful.

Despite the existence of multiple frameworks to evaluate aid effectiveness, such as the 2005 Paris Declaration [28], the Good Humanitarian Donorship Agenda [35] and the Fragile States Principles [36], there have been limited attempts to implement and adopt these standards, particularly in conflict-prone regions [14].

Similar to previous findings in this research consortia’s publications on aid alignment [37] and aid displacement [16], our literature review uncovered scant evidence of harmonisation of aid in conflict-affected areas with a notable absence of studies on the Eastern Mediterranean Region(EMR) region. Therefore, based on our best knowledge, this is the first paper that assesses aid harmonisation in a conflict-affected country in the EMR region.

This work will support donors in addressing the various political and logistical challenges of transitioning to early recovery and the challenges associated with inequitable aid following the earthquake in Syria in February 2023 [2] by using lessons learned on aid effectiveness over the past decade. Such a review would also be helpful to the broader region, considering the high number of protracted conflicts requiring long-term aid delivery.

The broader aim of this study is to assess aid effectiveness based on the pillar of harmonisation in the Syrian humanitarian response between 2011 and 2019 based on the Paris Declaration’s five principles (see Table 1 for key terms). The specific focus of this paper is aid harmonisation as defined by the Organisation for Economic Cooperation and Development (OECD) by analysing the trends in pooled funds and aid fragmentation, and their consistency with the key indicators of the Syrian conflict. The study also evaluates evidence of aid harmonisation of all donors in Syria by examining the percentage of the pooled fund in Syria relative to other fragile states and low and middle-income countries.

## 2. Methods

### 2.1. Study design

This is a mixed methods country case study that explores aid harmonisation in Syria between 2011 and 2019. The study design was informed by the OECD’s survey on monitoring the Paris Declaration [38], [39] and by a methodology for assessing aid effectiveness by Burall and Roodman in 2007 [40]. The study assesses aid harmonisation based on two indicators mentioned in “The Paris Declaration on Aid Effectiveness and the Accra Agenda for Action” [28]: pooled funds, which serve as a “joint financial arrangement” for donors to coordinate and increase funding efficiency and reduce aid fragmentation that leads to higher transaction costs for the recipient [41]

### 2.2. Quantitative Analysis

#### 2.2.1 Data sources

Data on humanitarian and health aid for Syria were collected from the OECD’s CRS [42]. This database is the most comprehensive for tracking health and humanitarian aid for conflict-affected countries; it allows analysis of different aid activities, multilateral and philanthropic donors, country donors and recipients, purpose of aid, policies, and years [43], [44]. In addition, it covers the specific economic or social programs that the aid aims to support in a recipient country and classifies these by sector. Also, some descriptive information on the projects is provided [45].

The data in the CRS are reported by 49 bilateral donors, 42 multilateral donors, and 36 private donors. The CRS provides financial data for 2002 -2021, with almost 200,000 to 300,000 data entries per year. Current limitations are that some donors, including China, Qatar, Saudi Arabia, and some private funding sources, do not report their aid disbursements to the system[44], [46]–[48]. Additionally, some donors, such as Turkey, do not accurately report their aid [14] However, we did not find any other comprehensive sources tracking aid in Syria.

CRS financial data were collected from 2011, with the onset of conflict in Syria, to 2019. The CRS data used in this study are based on the 31 April 2021 update [42] and were downloaded to Excel on 15 August 2021. The CRS code list was updated on 24 April 2021 [49]. We then utilised trend analysis techniques through Excel to track their progress, enabling us to understand the correlation between aid disbursements and crisis indicators. At the same time, we investigated the links between the timeline of the crisis and the changes in health and humanitarian aid trends.

We identified four key humanitarian indicators to explain the salient features of the conflict’s impact on the population inside Syria: 1) the number of IDPs; 2) the number of people in need of humanitarian assistance; 3) the number or frequency of internal movements (displacements) by individuals due to conflict; and 4) the decline in Syria’s population between 2011 and 2019 [14].

Figure 1 illustrates a stark downward trend in Syria’s population since 2011, with a decrease of approximately 4.7 million observed over the course of the study period. In contrast, the number of IDPs experienced a marked increase throughout the same period, with a peak in 2014 and further increases in 2017 and 2019. Internal movements also peaked in both 2013 and 2017, followed by two consecutive years of a spike of population needs in 2016 and 2017, with 13.5 million people in need in each of these years.

**Figure 1.**
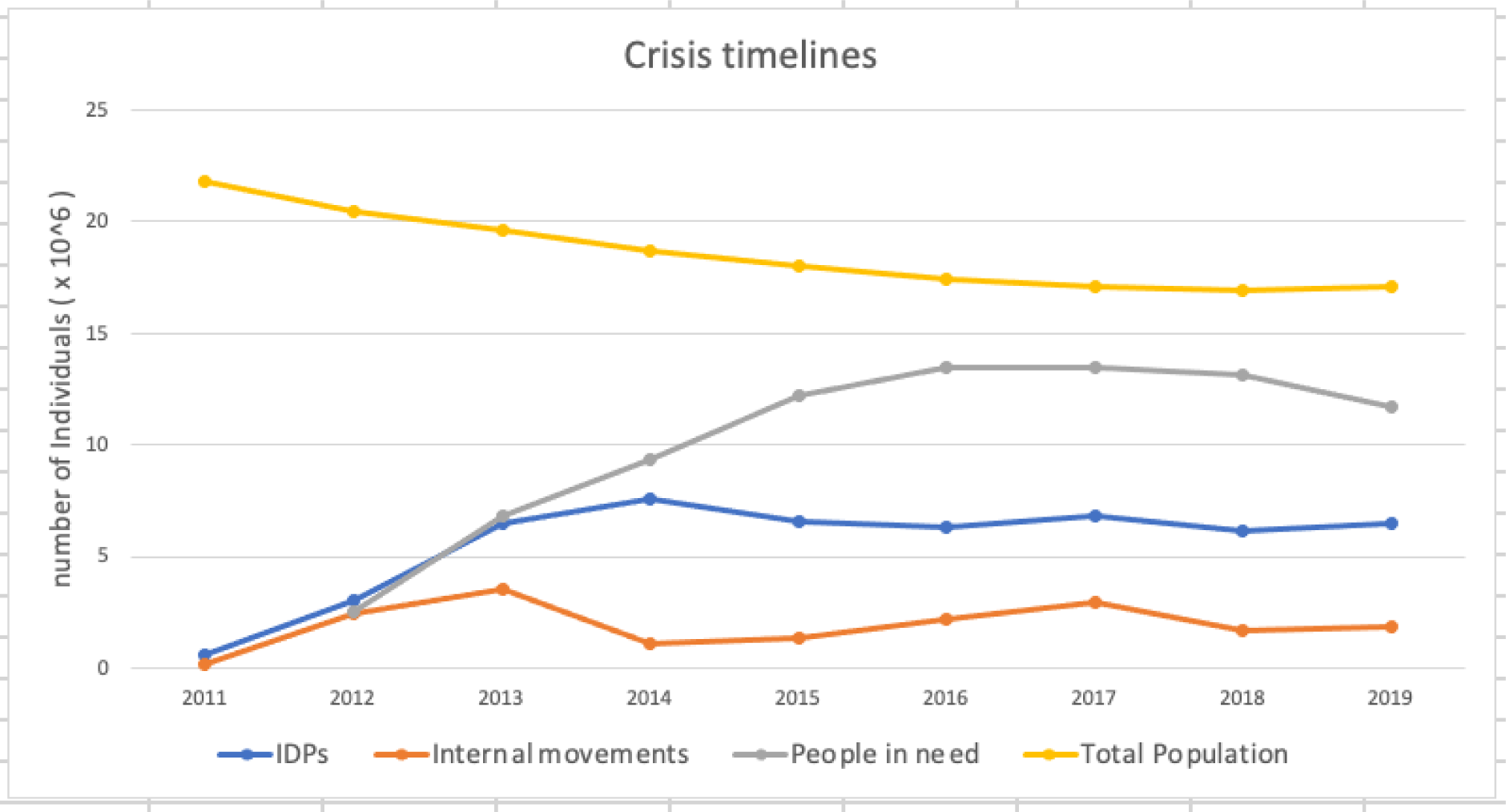
Crisis Timeline in Syria 2011-2019 [14]

The four indicators reflect the severity of the humanitarian crises throughout the study period. 2017 was the worst year due to the large number of violent incidents in 2016 and 2017 [50]. Following the Russian military intervention, starting in alliance with the Syrian regime in the autumn of 2015, the level of violence increased dramatically [51]. This resulted in 338 confirmed attacks on healthcare facilities across Syria killing dozens of healthcare workers in 2016 [50], evacuation of rebel-held east Aleppo in 2016 [52], bombing and besieging some 400,000 people in Eastern Ghouta [53], and the 2017 chemical attack on Khan Sheikhoun [54].

In this analysis, our definition of aid includes “Official Development Assistance” (ODA), “ODA grants”, “ODA loans”, and “Private Development Finance.” The definition excludes “Equity Investment” and “Other Official Flows” [42], [43], [55]. This is consistent with other recent studies [43]. Data on regional and non-country-specific aid were not included in this analysis, and the focus was solely on the flow of aid to the Syrian areas, and therefore does not include aid for Syrian refugees in neighbouring countries or overseas.

We extracted data on gross disbursements rather than commitments, as we were looking for “the actual international transfer of financial resources, or goods or services valued at the cost to the donor.” [56] To analyse aid trends over lengthy periods, we relied on constant 2019 US dollars rather than the current value to account for exchange rate fluctuations and inflation. The CRS aid database contains bilateral ODA of the Development Assistance Committee (DAC) members and excludes their contributions to the regular budgets of multilateral institutions when accounting for bilateral aid [57]. We did not include Turkey in our analysis due to a classification error in their reporting on the CRS system [14]. Turkey designates the aid costs of supporting Syrian refugees in Turkey as an ODA grant, even though these costs do not go to the people in Syria [14].

In the realm of industrial organisations, researchers frequently use specific indicators to navigate the complexities of competition. Among these, are the Herfindahl-Hirschman Index (HHI) and the concentration ratio (CR). The most frequently used indicator is the HHI, found in many recent studies to evaluate humanitarian aid fragmentation [33], [43]. We examined the fragmentation indicator using the HHI. The HHI is calculated by “squaring the market share of each competing firm in the market and then adding the resulting numbers [58].” When there are many donors giving similar amounts of aid, the index approaches close to zero (high level of fragmentation), while it reaches a maximum of 10,000 when only one donor is providing all aid (high level of concentration). The index increases as the variation in aid contributions increases and the number of donors decreases [58].

The scope of the study is the whole of Syria, therefore including government-controlled areas and non-government-controlled territories.

#### 2.2.2 Quantitative variables

Based on the abovementioned databases, we identified several donor disbursement variables representing pooled funds and aid fragmentation. The following table summarises the variables used in the quantitative analysis:

### 2.3. Qualitative analysis

To further our understanding of aid harmonisation in Syria, we complemented the quantitative analysis with four FGDs, four semi-structured interviews and three individual consultations with experts, public sector officials, and humanitarian practitioners. This was crucial as the quantitative data alone did not comprehensively reflect the local context and developments on the ground as the data demonstrates donors’ perspective on harmonisation rather than that of implementing organisations, local health authorities and beneficiaries.

We used purposive sampling followed by snowballing sampling approaches to identify the FGDs participants. The research team invited 31 senior humanitarian workers from medical NGOs and INGOs, local authorities, technical institutions, and the Gaziantep-based Health Cluster to attend the FGDs in Mersin, Turkey, in August 2021. Of the 31 invitees, 25 participated in the FGDs. Eighty-eight percent of the participants have medical backgrounds and extensive experience in humanitarian and health programs. Twenty percent practice their medical profession inside Syria in addition to their affiliation with NGOs and quasi-governmental institutions.

The distribution of participants was as follows: Ministry of Health in the Syrian Interim Government [2], Idlib Health Directorate [2], Health Information System Unit [1], Syrian Board of Medical Specialties [1], INGOs [6], NGOs [12], Health Cluster [1]. Four FGDs were conducted, two at a time, with an average of 12 participants for each group. We ensured all groups had equal representation and sound balance of stakeholders during the FGDs.

Discussions were conducted in Arabic, with a moderator and note-taker for each group. FGDs were not recorded, and ethical consent, including disseminating and publishing the study findings, was taken verbally, based on participant preference. The results were presented in an aggregated and anonymised format to protect participant privacy.

The FGDs were followed by four KIIs with representatives from four leading donors in Syria involved in the humanitarian and health response in various areas of control, including areas under the Syrian regime control. The interviews aimed to validate our results and initiate policy implications. The feedback from these semi-structured interviews was consistent with the areas discussed in the FGDs. The KIIs were conducted in English and recorded. They were later transcribed and anonymised using a unique identifier for each participant. Following thematic analysis, data from the FGDs and the KIIs were extracted and categorised into different themes. Written ethical consent from the participants in the KIIs was obtained.

Finally, we discussed the recommendations through individual consultations with experts in the humanitarian and private sectors in Syria. Ethical consent was taken verbally.

## 3. Results

### 3.1. Trends in humanitarian and health pooled fund flows concerning the key conflict and crises indicators

Figure 2 shows the crisis indicators and the aid percentages as a pooled fund for health and humanitarian aid. It shows a peak in pooled health aid (purple dashed line) to 0.6% in 2013. This corresponds to the rise of the internal movement indicator. However, no correlation with crisis indicators was observed outside of 2013, with the percentage remaining close to zero for the duration of the years studied. From 2016 to 2019, there was a marked increase in pooled humanitarian aid among donors, as indicated by the increase in the green dashed line in Figure 2.

**Figure 2.**
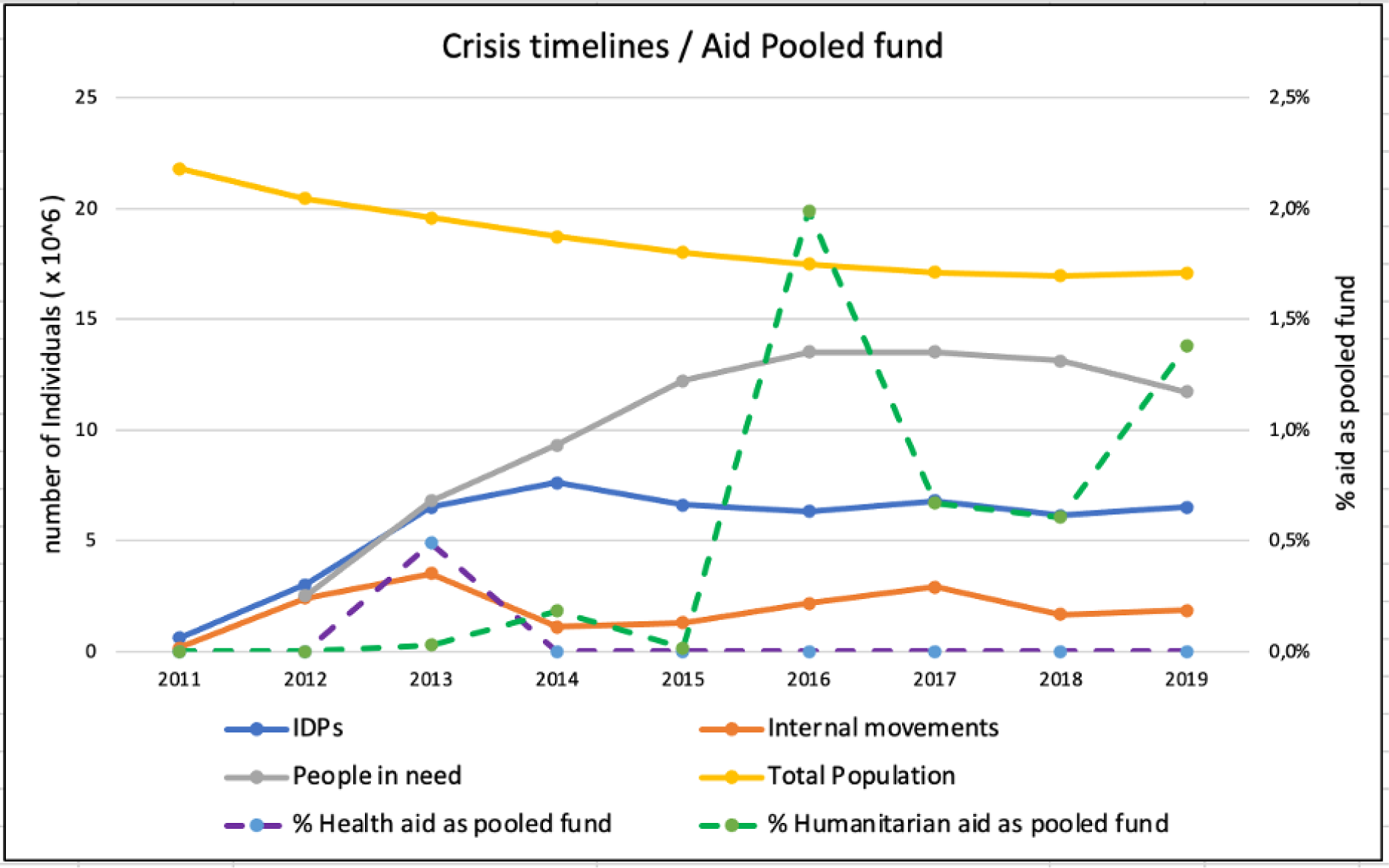
Percentage of health and humanitarian aid as pooled fund concerning the key crises indicators, 2011-2019.

According to focus group participants, aid harmonisation in Syria was not significantly affected by changes on the ground or needs assessments in the health and humanitarian sectors. Instead, it was the WoS coordination mechanism which had the most significant impact on harmonisation.

### 3.2. Health and humanitarian aid as pooled fund compared to other low and middle-income countries and fragile states

Figure 3 shows that the proportion of health pooled funds in Syria is almost non-existent, with a slight increase in 2013, when compared to the 1-5% observed in low- and middle-income countries and highly fragile states. Interviewees cited the reluctance and the lack of interest of donors to invest in health aid for development in Syria as a potential reason behind the very low proportion of pooled funds. Furthermore, since health funds were limited, donors have not made a concerted effort to coordinate their activities or implement joint projects in Syria.

**Figure 3.**
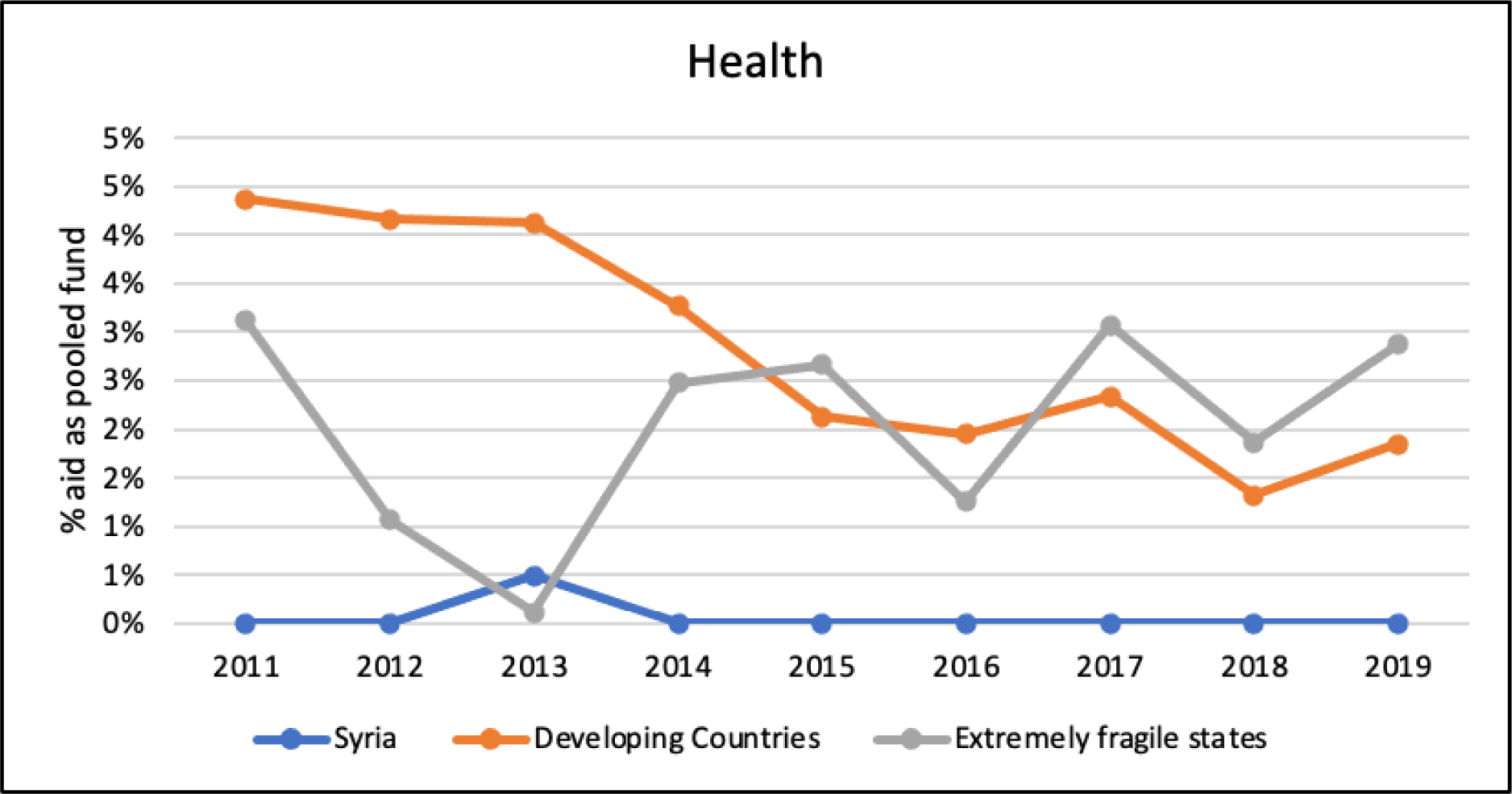
shows the percentage of health aid as pooled funds (2011-2019) in Syria, LMICs and extremely fragile states (CRS).

In contrast to health aid, it can be observed from Figure 4 that the proportion of the pooled fund in the humanitarian sector in Syria over the first five years of the conflict was close to negligible and significantly lower than in other fragile and low- and middle-income countries. However, in 2016, there was a noticeable increase, with the highest estimated rate being 2%. This trend continued for the following three years, with rates ranging between 1% and 2%. This indicates an improvement in the harmonisation of donor countries’ efforts in the humanitarian sector in Syria from 2016 to 2019.

**Figure 4.**
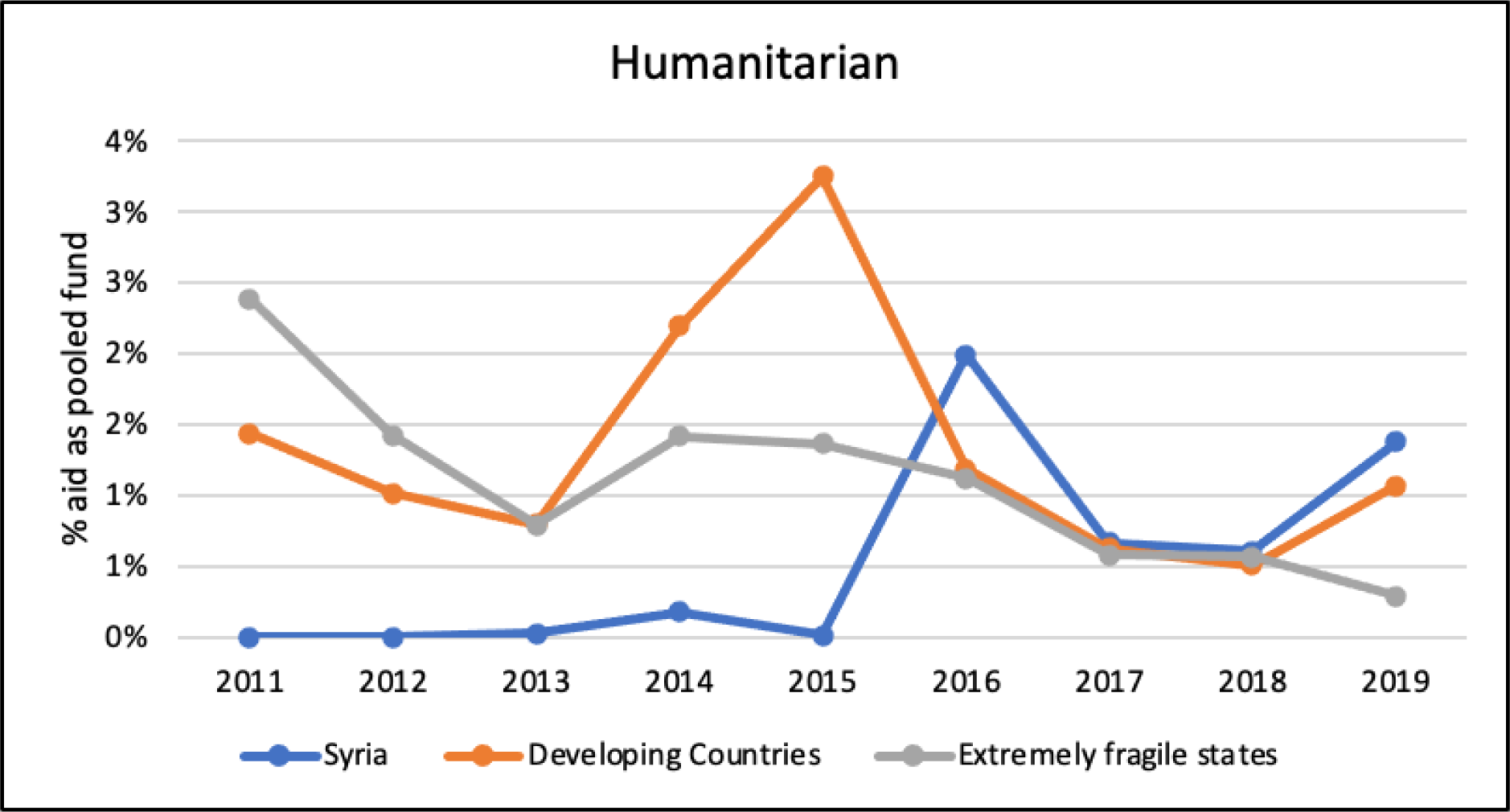
Percentage of humanitarian aid as pooled funds 2011-2019 (CRS).

Drawing from the participants’ insights, pooling funds from multiple donors could be a challenging endeavour, as the interests and views of donors often differ, and the Syrian context is highly complex. Nonetheless, the protracted crisis in Syria and the vast amount of humanitarian assistance provided has enabled donors to establish coordination mechanisms, such as the WoS approach in 2015. It was noted that the proportion of pooled funds for cross-border humanitarian programs in Syria is among the highest in the world, and that the country-based humanitarian pooled fund has a considerable percentage allocated to health programs.

### 3.3. Humanitarian and health aid fragmentation

The fragmentation index of humanitarian and health funding during the study period ranged from approximately 1500 to just below 3000, as can be seen in Figure 5. This indicates a moderate concentration of aid during the study period. While there has been a general trend towards increased fragmentation of humanitarian aid over time, health aid fragmentation has been more volatile, particularly in 2013.

**Figure 5.**
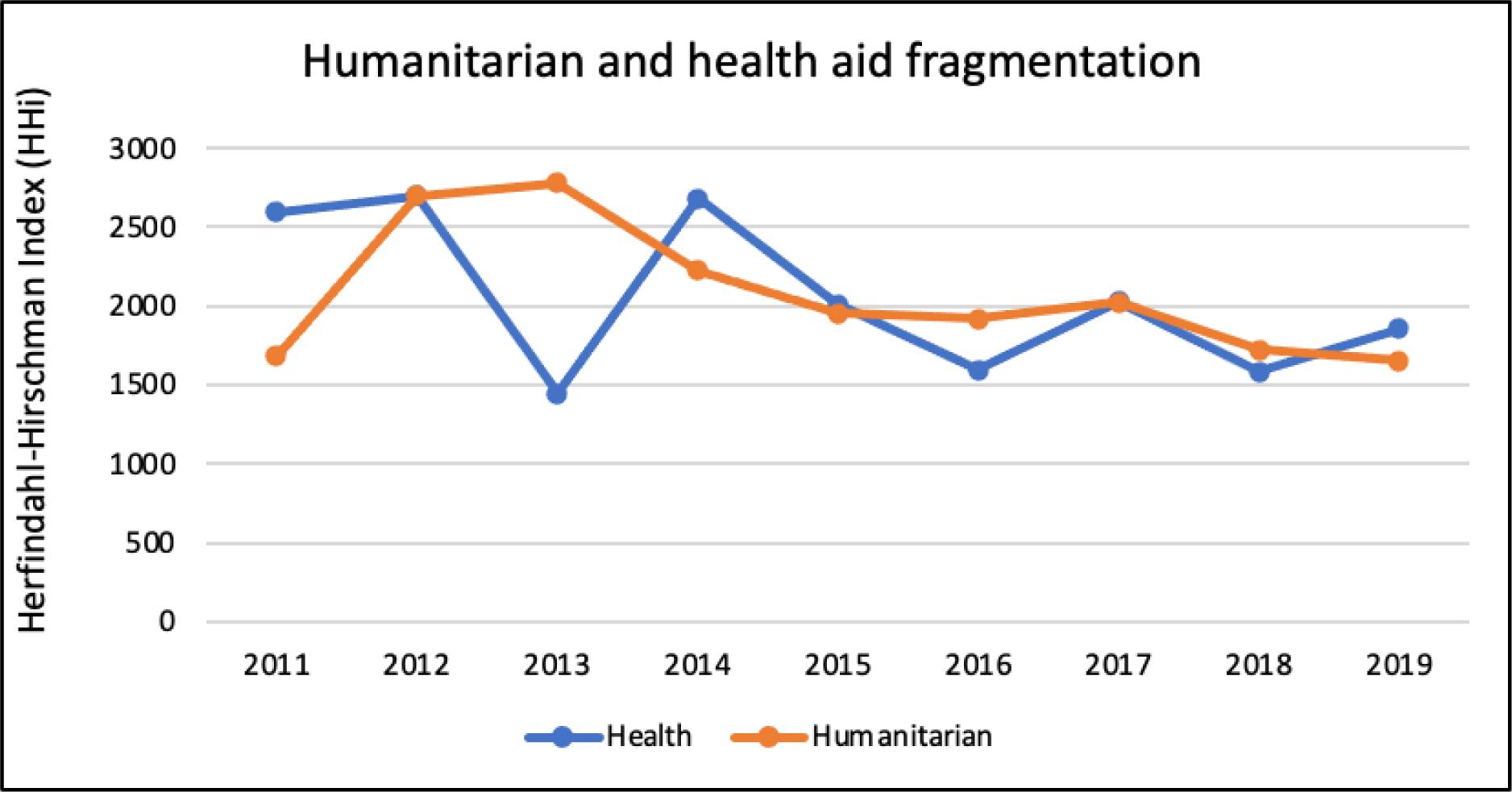
Humanitarian and health aid fragmentation in Syria 2011-2019 (CRS).

Many participants questioned the concept of fragmentation as an indicator of aid effectiveness, arguing that, for certain sectors, a limited number of coherent donors could be beneficial for stabilisation and sustainability. Even though, it could be a risk factor in other sectors and crises. Most participants agreed that the reality of fragmentation is likely to be more pronounced than the figures indicated because once programmes go to the level of implementing agencies, further fragmentation appears, even if the programme is supported by pooled funds. Consequently, it was suggested that the indicator should be studied at the levels of contracting channels and implementing agencies, not just at the donor level. Moreover, there was a consensus that a new indicator should be introduced to help elucidate the actual impact of fragmentation on the health sector. This is because a greater number of donors may be necessary to prevent significant funding gaps, due to the complexity of the context and the risk of any donor withdrawing support at any time. While some participants considered some sectors that lack funding, such as health, might benefit from having fewer donors because this would ultimately increase funding efficiency, although the risk of depending on fewer donors remains.

## 4. Discussion

The OECD has developed several frameworks and indicators to assess the various elements of aid effectiveness; pooled funds and aid fragmentation [28] have been identified to investigate aid harmonisation in this study. These have been explored in other contexts, including Bangladesh [13], Mozambique [60] and Vietnam [61]. Using the framework of the Paris Declaration, our results examined aid harmonisation throughout nine years of the Syrian conflict. Our study found clear trends regarding pooled funds and aid fragmentation and gaps that can be improved with regard to aid harmonisation in this highly complex protracted conflict-affected context.

First, aid harmonisation did not occur in Syria and importantly it did not correlate with increased humanitarian needs and is not conflict sensitive. Mark Lowcock, the coordinator of the UN’s aid relief operation, said in 2021, “The humanitarian system is set up to give people in need what international agencies and donors think is best, and what we have to offer, rather than giving people what they themselves say they most need” [62]. These practices contradict what donors are expected to do in humanitarian contexts, where decisions relating to aid allocation and mechanisms should be based on both local requirements and national systems’ performance [63].

In conflict zones, humanitarian policies face a changing geopolitical environment in which traditional humanitarian norms face wavering support [64]. A study conducted in Indonesia showed correlations between the lack of harmonisation and the pathologies of foreign aid, such as the prevalence of certain donors’ strategic interests and a bureaucratic incentive structure, rather than a lack of government coordination [65]. A study in Ghana accentuated the value of integrating beneficiary views on the impact of projects first in the implementation stage instead of waiting until the end of the project. Donors should, therefore, adjust their expectations to fit those of the beneficiaries [66]. Many studies in Syria about the COVID-19 and earthquake responses emphasised the importance of bottom-up governance and community engagement approaches to enhance aid effectiveness [67]–[70].

The politicisation of aid has had a significant impact on aid harmonisation over the study period (2011 – 2019). However, an increasingly negative impact occurred after the closure of three out of four cross-border crossings for UN-governed humanitarian aid in 2020 [71]. These closures were due to the vetoes of Russia and China in the Security Council. Furthermore, in July 2023, Russia’s veto on the UN Security Council blocked further imports of UN aid into northwest Syria, partially shifting control to the Syrian regime [72]. Nonetheless, the UN was authorised by the Syrian regime to use Bab al-Hawa, the last approved crossing for UN aid, for another six months starting on 13 July [73]. This situation paved the way for what is known today as ‘The consent model’ [74], leaving 4.5 million people in northwest Syria [75] facing uncertainty regarding UN aid for the area and putting them at risk of aid weaponisation [76] by the Syrian regime.

The WoS coordination architecture recognises the NES NGO Forum as a response modality for NGOs operating in northeast Syria [77]. However, since there is no Security Council Resolution or approval from the Syrian regime to use the Al Yaroubieh crossing, which was closed in 2020 [71], the role of the NES Forum is limited to coordinating aid within the area and cross-line aid. However, most INGOs supporting northeast Syria are based in Erbil, Iraq, which causes a coordination gap between INGOs, NGOs and UN agencies. Additionally, the absence of UN cross-border aid to northeast Syria has negatively impacted humanitarian funds for 2.7 million people [75], [78], [79] and thus humanitarian pooled funds.

Second, evidence from other LMICs emphasises the need for collaboration to coordinate activities between organisations, especially in situations where objectives are complex and the health and humanitarian needs are immense [80]. Despite several international declarations and efforts to make aid more effective for beneficiaries, this study shows that there is significant room for improvement in all these mechanisms, especially in conflict-affected contexts. Several models and approaches for aid effectiveness and allocation aim to ensure aid harmonisation, including Performance Based Allocation (PBA) models, models emphasising national (and local) prerequisites, approaches based on the selection of ‘focus’ countries or pre-selection mechanisms, and demand-driven approaches. Allocation of funds should support the highest-priority projects of the best-placed responders through a transparent and inclusive process that supports the priorities of Humanitarian Response Plans (HRPs). This ensures that funding is available and prioritised locally by those closest to the affected population [81].

Third, during the first five years of the conflict (2011-2015), humanitarian and health pooled funds (which endorse aid harmonisation) in Syria were nearly entirely absent, far less than those in LMICs and highly fragile states. Pooled funds have evoked an abundance of recent discussion, yet concrete evidence of the effectiveness of these is required. The 2011 World Development Report stated that pooled funds have had mixed results, with criticisms including inadequate expectation management and slow outcomes due to difficulties in working through national systems [82]. In addition, there is a scarcity of relatively comparable data on pooled fund performance, making it challenging for donors, recipients, and implementers to evaluate the efficiency of this mechanism [83]. Moreover, no agreed upon system or tool can be utilised to examine pooled funds [84].

From 2016 to 2019, a visible surge in humanitarian pooled funds indicated an increase in the harmonisation of donors’ efforts influenced mainly by adopting the WoS approach in 2015 according to the interviewees. Since the WoS approach is the only known mechanism to synchronise donors’ activities during that period. Furthermore, the participants noted that many donors favour contributing to multilateral funds, such as pooled funds. These are perceived as a sharing of risk among donors for corruption and facilitate more rigorous and diligent reporting. This may explain the improved pooled humanitarian fund in Syria after 2016, to address the increased risks that stemmed from the rising complexity of the conflict. Nevertheless, most donors still opt for bilateral contracting due to their respective laws or strategic plans, such as funding the local Red Cross/Red Crescent activities and INGOs.

WoS was developed to streamline humanitarian efforts in Syria and across its borders and to overcome inefficiencies, gaps in services, and ensure strategic and operational coherence among different stakeholders [85], [86]. It covers various sectors, such as health, education, food security, water and sanitation, shelter, protection, and early recovery [87]. Furthermore, it brings together over 270 national and international actors to coordinate aid logistics, governance mechanisms, and resource allocation [85]. The WoS was emerged as a positive result to the cross-border UNSCR in 2014.

According to the participants in the FGDs, the WoS approach was essential for life-saving services, as it provided a comprehensive framework to address urgent health needs, from primary care to emergency medical services. It attempted to bring consistency to aid effectiveness, governance, and the legitimacy of humanitarian actions, aligning with global health goals even in the challenging context of conflict. This coordinated model has been subject to analysis in this paper for its effectiveness in achieving a more organised and impactful humanitarian response.

Participants also believe that this mechanism was introduced by donors partly to minimise the risks to the donors associated with working within the increasingly complex Syrian context, especially as sanctions and political redlines imposed by donors’ countries made it increasingly difficult to deliver humanitarian aid.

The reason behind almost absence of the (development) health pooled fund over the study period seems to be the reluctance of donors to invest in health infrastructure and governance, which would have meant investing in the public health system in government-controlled areas where a long list of sanctions [88] have been imposed on the Syrian regime, involved in many war crimes [89]. Additionally, due to the political readiness of donors. Some informants also think this is because Syria was not seen to have entered a stable post-conflict phase. Donors, therefore, favoured supporting humanitarian health aid under the humanitarian funds’ umbrella. Our previous study on aid displacement demonstrated that between 2011 and 2019, the total amount of funding allocated towards humanitarian aid was fifty times greater than towards (development) health aid, displaying a notable preference by donors to put money into the humanitarian sector compared to the health sector [90]. This preference only creates further difficulty in transitioning to early recovery.

Fourth, no evident financial fragmentation has been found over the study period at the donors’ level. However, the real fragmentation exists at the implementing partners’ level, according to the participants. Additionally, the health system in Syria has commonly been described as a fragmented health system [69].

Improved division of labour can lead to a concentration of donors focusing on specific countries or sectors, including health or humanitarian, which can in turn decrease transaction costs and facilitate coordinated efforts. This approach can also guarantee comprehensive coverage for all countries, rather than only those favoured by donors [61]. Nevertheless, the effect of fragmentation on aid efficiency has an equivocal effect that is determined by the sector and level that the aid is administered. Studies suggest that fragmentation in aid lowers economic growth, resulting in increased corruption and a decrease in technical assistance. In contrast, recent research implies that it can positively influence child survival and the promotion of democracy [91]. These confounding results can be due to the variations between sectors and the level of aid implementation across different areas. To address this issue, the specific contexts [92] and the local aid strategies must be considered to direct donors’ policies regarding financial aid fragmentation.

Regardless of its impact, aid fragmentation exists when considering it from the perspective of developing countries, as many donors give relatively small amounts of aid. According to the OECD survey report from 2005 to 2006, Vietnam had 29 major donors, of which 17 made up solely 10% of the assistance it received [61], while in 2009 Mozambique had 12 minor aid donors, which signalled significant aid fragmentation [93].

## 5. Recommendations

To improve aid effectiveness, donors should seek to improve an aid strategy model in conflict settings by considering public health authorities’ legitimacy, power dynamics, performance, health governance principles, humanitarian needs, and community-based, rights-based, and conflict-sensitive actions. Additionally, this strategy should be based on a negotiation process between donors and parties involved in a conflict. Nonetheless, fragile contexts exhibit widely varying characteristics, and the opportunities are very particular, with instruments affecting each context uniquely. Thus, development actors should be wary of ‘one size fits all’ strategies [92].

In Syria, donors should apply an adaptable funding strategy to meet the early recovery requirements, including multi-year funding and risk sharing with local Syrian NGOs (not risk transfer) and a community-centred aid approach. This strategy can be implemented according to the readiness of each area of control: northwest, northeast and GoS-controlled areas.

Our results show that there is a need for an improved examination of aid effectiveness in humanitarian settings to include the perspective of the local community and assess the ability of aid to be responsive to the actual humanitarian needs rather than other considerations, such as the political ones. The 2005 Paris Declaration acts as a platform developed primarily by the donor community, thus emphasising the significance of establishing frameworks and tools to analyse aid effectiveness both from donor and beneficiary perspectives.

Harmonisation mechanisms must be disentangled from polarised international politics to improve aid effectiveness as much as possible. One of the solutions that emerged in 2022 is the Aid Fund for Northern Syria (AFNS) initiative, a humanitarian multi-donor pooled fund established in Gaziantep, Turkey, to address priority needs in northern Syria with a primary focus on the northwest [94]. This platform does not require a Security Council Resolution to manage cross-border humanitarian aid. Such an approach should be evaluated, and its activities should be expanded to cover northeast Syria in collaboration with the NES NGO Forum. This approach can increase pooled fund and offers more sustainable health and humanitarian responses. However, AFNS is currently only able to cover gaps that will be left in case ‘The Syria Cross-border Humanitarian Fund (SCHF)’ is stopped. So, AFNS’ funds should be increased, and its policies should not repeat the WoS and UN clusters; instead, they should include a development and localisation agenda to deal with the protracted compound crisis in Syria and the early recovery requirements.

There is uncertainty about the fate of the Gaziantep Hub, which is the main coordination platform for humanitarian aid in northwest Syria, as the Syrian regime’s approval for the UN-governed cross-border aid expires in December 2023. Additionally, ‘The consent model’ is not an acceptable model from the local actors [95] and communities [96] and is not a sustainable one. Therefore, an alternative coordination mechanism or platform should be urgently prepared. This platform should be conflict-sensitive and guarantee the continuation of cross-border aid.

## 6. Limitations

- We did not include casualty figures in the crisis timeline. Although it is a significant indicator, no reliable casualty data covers the duration of the study. The UN ceased its official count of fatalities in Syria by January 2014.
- We excluded 2020 and 2021 data from our analysis as it was yet to be available on the CRS when we downloaded the data for analysis and validation through FGDs and KIIs.
- Most of the aid missions facilitated by the specialists in the FGDs occur in northern Syria, with limited access to areas under the control of the Syrian regime.

## 7. Conclusion

This study highlights the complex landscape of aid harmonisation in the Syrian crisis. It provides a new in-depth examination of the Paris Declaration framework and how it could be improved. Notably, overall aid harmonisation in Syria did not occur and importantly did not correlate with increased humanitarian needs and crisis indicators over the study period.

The surge in humanitarian pooled funds after 2015 suggests an improvement in donor harmonisation, primarily attributed to the WoS approach, which emerged as a positive result of UNSCR in 2014. No evidence of aid fragmentation at the donor level has been found. However, the fragmentation at the implementing partner level may present a different perspective.

The research concludes that aid harmonisation in humanitarian settings should be assessed using approaches incorporating community engagement and end-user perspectives.

The study also calls for alternative mechanisms and platforms for cross-border humanitarian operations to avoid the negative impact of politicising aid decisions by the Syrian regime and Security Council.

## Declarations

## Acknowledgement

We would like to express our gratitude to all the participants who took part in the focus group discussions and key informant interviews. We would also like to extend a special thanks to Mr Fadi Al-Dairi from Hand in Hand For Aid and Development organisation and Dr Catherine Smallwood for reviewing the last version.

## Authors’ contributions

The initial conceptual framing, literature review, focus group discussions (FGDs), key informant interviews (KIIs), initial drafting of the piece, multiple rounds of edits, and producing the final manuscript were carried out by MAK. AE, MAA, and ZAZ contributed to data collection and data analysis. All the authors contributed to further literature review, additional content, and a round of edits. KM and PP contributed to the overall structuring, analytical content and producing the final draft. All authors read, edited and approved the manuscript.

## Reflexivity Statement

Many authors of this paper are Syrians who have been involved in the health and humanitarian responses to the Syrian humanitarian crisis since 2011. The lead author Dr Munzer Alkhalil co-founded many health qazi governmental institutions in northwest Syria, including the Idlib Health Directorate (IHD); he was the head of IHD between 2011 and 2020. As such, Syrian authors bring direct experience and understanding of humanitarian response and mechanisms in Syria. International academic authors also guaranteed the balance between local and global perspectives and practical and theoretical dimensions. Finally, this paper is the third one related to this research consortium’s series on aid effectiveness, following previous works on alignment and aid displacement.

## Availability of data and materials

The datasets generated and analysed through the FGDs and KIIs are not publicly available as they contain information that could compromise research participant privacy. All other data sources used in this study were publicly accessible.

## Ethical approval and consent to participate

Ethical approval to conduct the FGDs was obtained from the Idlib Health Directorate in northwest Syria (16 June 2021, reference number 2307). Additional ethical approval for the KIIs, was obtained from King’s College London (22 September 2021; MRA-21/22-26339). The study has ensured that all quotes of interviewees remain anonymous.

## Consent for publication

Verbal consent was obtained from participants in the FGDs and individual consultations, and written consent was obtained from participants in the KIIs. Participants were informed that the findings might be shared and published in academic journals, conferences, or other scientific platforms. Their personal information and identity remain confidential, and all necessary steps were taken to ensure anonymity and privacy. The results were presented in an aggregated and anonymised format to protect participant privacy. By providing consent, participants agreed to disseminate and publish study findings while ensuring confidentiality and privacy.

## Patient consent for publication

Not applicable.

